# Machine learning-supported interpretation of kidney graft elementary lesions in combination with clinical data

**DOI:** 10.1101/2021.09.17.21263552

**Authors:** Marc Labriffe, Jean-Baptiste Woillard, Wilfried Gwinner, Jan-Hinrich Braesen, Dany Anglicheau, Marion Rabant, Priyanka Koshy, Maarten Naesens, Pierre Marquet

## Abstract

**Background:** The Banff classification standardizes the diagnoses of kidney transplant rejection based on histological criteria. Clinical decisions are generally made after integration of the Banff diagnoses in the clinical context. However, interpretation of the biopsy cases is still heterogeneous among pathologists or clinicians. Machine Learning (ML) algorithms may be trained from expertly assessed cases to provide clinical decision support.

**Methods:** The ML technique of Extreme Gradient Boosting learned from two large training datasets from the European programs BIOMARGIN and ROCKET (n= 631 and 304), in which biopsies were read centrally and consensually interpreted by a group of experts and used as a reference for untargeted biomarker screenings. The model was then externally validated in three independent datasets (n= 3744, 589 and 360).

**Results:** In the three validation datasets, the algorithm yielded a ROC curve AUC of mean (95% CI) 0.97 (0.92-1.00), 0.97 (0.96-0.97) and 0.95 (0.93-0.97) for antibody-mediated rejection (ABMR); 0.94 (0.91-0.96), 0.94 (0.92-0.95) and 0.91 (0.88-0.95) for T cell-mediated rejection; >0.96 (0.90-1.00) in all three for interstitial fibrosis - tubular atrophy (IFTA). Finally, using the largest validation cohort, we developed an additional algorithm to discriminate active and chronic active ABMR with an accuracy of 0.95.

**Conclusion:** We built an Artificial Intelligence algorithm able to interpret histological lesions together with a few routine clinical data with very high sensitivity and specificity. This algorithm should be useful in routine or clinical trials to help pathologists and clinicians and increase biopsy interpretation homogeneity.

## Introduction

The international Banff classification standardizes the diagnosis of different forms of kidney allograft rejection.^1^ It is based on a grid of histological criteria with predefined thresholds, ranking the extent of elementary lesions. Subsequently, numerous rules must be applied on the possible combinations of histological lesions to deduce: antibody-mediated rejection (ABMR); T cell-mediated rejection (TCMR); and others, including interstitial fibrosis and tubular atrophy (IFTA, that is no longer considered as a category in itself).^1^ However, this gold-standard strategy is not perfect and the transcription of elementary lesions into a final clinical interpretation may appear challenging. First, the interobserver reproducibility of reporting and ranking histological lesions is suboptimal.^2–4^ In a recent international survey, 6 case-based scenarios in which the elementary Banff lesions and clinical background were provided (i.e. the slide reading step was skipped), were interpreted by 95 clinicians and 72 renal pathologists. Case interpretations differed by 26 and 34% from the reference standards, respectively.^5^ The absence of DSA or negative C4d staining, or serum creatinine, proteinuria and other clinical data may influence the pathologist’s or clinician’s judgement of a case. Secondly, the definition of the phenotypes has dynamically evolved since 2005 with each revision of the Banff classification, leading to changes in the evaluation criteria applied and in the diagnostic entities, including their designation by name.^1^ In small transplant centers where pathologists have a general practice, it is challenging to integrate these frequent updates with the same level of expertise as pathologists specialized in the analysis of kidney allograft biopsies. Thirdly, centralized assessment and consenting on biopsy diagnoses by specialized pathologists is not always possible in clinical studies in kidney transplantation (e.g., on biomarkers,^6^ treatment strategies, survival analyses in prospective or retrospective cohorts) due to logistic and financial constraints.

Three strategies may be used for the assessment of humoral rejection, t-cell rejection, or other biopsy lesions. The first is to apply the Banff rules strictly and automatically with “if then else” rules. The second is to identify clusters of elementary lesions in an unsupervised manner that cannot be directly compared with the Banff reference classes. Their pertinence is generally evaluated based on further patient outcome, and logically clusters of cases with lesions have poorer survival than those with normal biopsies.^7^ The third strategy, as yet unexplored, is to identify case clusters in a supervised manner based on reference diagnoses made considering both the Banff classification and the clinical context, so as to automate and homogenize the process of clinical interpretation of biopsy cases used for clinical decision making.

Machine Learning (ML) is defined as a subset of the artificial intelligence (AI) domain, capable of automatically learning and continuously adapting interpretation or prediction algorithms. Robust mathematical procedures are applied by computer systems to achieve these complex tasks. With sufficient data, it can handle noisy and correlated variables, sometimes without the need for parametric assumptions, contrary to most traditional statistics. As recognized at the last Banff Meeting,^1^ the combination of quality and quantity of input data is key for achieving result quality using ML. Whatever the ML method, it is therefore necessary to train the model on a large enough database of pathological cases, examined by a panel of experienced pathologists.

The aim of the present study was to build a robust and accurate ML algorithm based on two large databases of biopsy cases interpreted by an expert group of pathologists and transplant physicians as part of two large European research programs. This algorithm was built to identify and hierarchize the Banff criteria and clinical data actually used by pathologists and clinicians to diagnose graft rejection, and repeat this process in a perfectly reproducible manner so as assign the exact clinical diagnoses to each biopsy in three large external datasets from various European countries.

## Methods

### Patients and biopsies

Histological data from kidney graft biopsies came from different independent datasets, in the form of the elementary Banff scores and reference diagnoses, as interpreted by pathologists and transplant physicians. For the training set, we used biopsy data from two European programs, BIOMArkers of Renal Graft INjuries (BIOMARGIN, ClinicalTrials.gov, number NCT02832661) and Reclassification using OmiCs integration in KidnEy Transplantation (ROCKET, funded by ERACoSysMed 2018-2021), both aiming at discovering and validating robust non-invasive biomarkers.^8^ The first two steps of BIOMARGIN were case-control studies enabling the untargeted search and then the selection of a broad list of biomarkers. The third, cross-sectional step aimed to validate the diagnostic performance of the biomarker candidates on a representative sample of transplant patients in Europe. Between June 2011 and August 2016, more than 650 sample triplets (urine, blood and biopsy) were collected in highly standardized conditions and stored in the Biobanks of the four hospitals participating in the project (Hôpital Necker Paris, France; University Hospitals Leuven, Belgium; Medizinische Hochschule Hannover, Germany; and Centre Hospitalier Universitaire Limoges, France). All these biopsies were read and interpreted locally and then sent for central reading by an independent expert pathologist, with adjudication of discrepancies by consensus between three independent expert pathologists. The final clinical diagnosis was made by four transplant physicians based on the consent histological interpretation and the clinical context.

Biopsy and omics data are still being gathered in our consortium as part of the ROCKET program, to discover accurate biomarkers of rarer phenotypes or graft lesions, including: active ABMR, chronic active ABMR, acute TCMR, chronic active TCMR, polyomavirus nephropathy (PVN) and glomerulonephritis. Ambiguous cases or those with confounding conditions and lesions are excluded. The corresponding dataset was used in the present study to train a more complex model able to distinguish active from chronic active ABMR. We could not study chronic inactive ABMR because the history of the cases was not available.

For the external validation of the ML algorithm and the choice of thresholds, we first used biopsy data from patients transplanted between 2004 and 2013 and followed-up until September 2019 at KU Leuven, Belgium. The second validation dataset was from patients followed-up from 2013 to 2019 at the Medizinische Hochschule Hannover, Germany and the third from a single-center study at Hôpital Necker, Paris, France,^9^ approved by the ethics committee of Ile-de-France XI (13016), where clinically-indicated renal allograft biopsies were collected from February 2011 to February 2013. All the patients of the external validation cohorts were different from those included in the above-mentioned BIOMARGIN and ROCKET studies.

For each biopsy, expert renal pathologists evaluated the elementary Banff criteria as recommended in the 2013 revised Banff Classification:^10,11^ glomerulitis (g), peritubular capillaritis (ptc), linear C4d staining in ptc or medullary vasa recta (C4d), chronic transplant glomerulopathy (cg), endarteritis (intimal arteritis, v), inflammation in non-scarred cortex (i), tubulitis in cortical tubules within non-scarred cortex (t), total cortical inflammation (ti), tubular atrophy in cortex (ct), interstitial fibrosis in cortex (ci), arteriolar hyalinosis (ah), arterial intimal fibrosis (fibrointimal thickening, cv). The diagnoses of interest were: active ABMR (yes/ no), TCMR (yes/no, borderline cases included as yes), IFTA lesions (grade ≥ II). These diagnoses were considered as the reference (*gold standard*) for training our ML algorithms. The clinical databases included the laboratory test results about donor-specific antibodies (DSA), serum creatinine (μmol/L) and proteinuria (g/L) at the time of the biopsy. No algorithm was built for glomerulonephritis or PVN as there were too few cases in the ROCKET training dataset and in the external validation datasets. Moreover, PVN is easily diagnosed by means of the specific SV40 staining, which is generally used in cases with positive BK virus serology. However, a computer program overlay was applied to biopsies with positive BK viremia together with positive t and i criteria, so as to avoid false positive TCMR diagnoses due to PVN.

### Statistical analyses

The predictors were: the Banff criteria semi-quantitatively scored from 0 to 3 (g, ptc, C4d, cg, v, i, t, ti, ct, ci, ah, cv); DSA positivity; serum creatinine, proteinuria, and time elapsed between transplantation and biopsy. Using the training dataset, a ML algorithm was built for each different outcome: active ABMR (yes/no), TCMR (yes/no), IFTA (yes/no) and ABMR (active/chronic active). In the training dataset, biopsies with more than 2 missing data among the elementary Banff lesions were removed. This exclusion was not applied in the validation datasets, so as to evaluate the algorithms in real-life situations. After analyzing the distribution of the Banff elementary lesion scores, we chose to impute the respective median value to the missing scores, in the training dataset. No imputation was made in the different validation datasets. The ML method of Extreme Gradient Boosting, an ensemble method based on decision trees, was chosen for its good performance on structured tabular data and its ability to handle missing data for making predictions.^12,13^ Prior to training the algorithms, we optimized the hyperparameters using ten-fold cross validation, for best accuracy. With this optimal set of hyperparameters (Supplemental Table 2), we assessed the algorithm performance in the training phase using the same ten-fold cross validation procedure. Receiver operating characteristic (ROC) curves, representing the true positive rate (sensitivity) vs. the false positive rate (1 - specificity), were used to assess the threshold-independent classification performance of each model. As the training dataset was imbalanced (skewed towards normal biopsies), we also used precision-recall (PR) curves^14,15^ representing precision (positive predictive value) vs. recall (sensitivity), not considering true negatives. The minimum PR area under the curve (AUC) is equal to the prevalence of the disease. When thresholds were set to a certain value, agreement between ML classification results and the expert conclusion was assessed by calculation of the accuracy. Accuracy is the probability that an observation is correctly classified (number of true positives plus number of true negatives, divided by the total number of individuals). In this study, the primary end points were the diagnostic accuracy and the ROC AUC of the different ML algorithms.

The Leuven cohort was used to set thresholds based on the accuracy, positive predictive and negative predictive values in this cohort.^16^ For external validation of the ML algorithms, these thresholds were then applied in the Hannover and Paris Necker cohorts.

For statistical computing and graphics, we used the free software environment R (version 4.0.3) and in particular, the xgboost package for classification (version 1.2.0.1).

## Results

In the BIOMARGIN training dataset (n = 643), 12 biopsies were excluded because they missed three or more Banff elementary lesion scores. Among the remaining 631 cases, 73 biopsies missed one Banff elementary lesion score and 29 missed two. Patient characteristics at the time of allograft biopsy are presented in Table 1 and other characteristics of the training dataset are detailed in Supplemental Table 1. Among the 304 biopsies of the ROCKET dataset, none had missing data (as it was a study exclusion criterion), 63 cases had active ABMR and 44 chronic active ABMR.

**Table 1.** Patient characteristics, laboratory test results at the time of allograft biopsy and histological diagnoses.

| **Variables** | **BIOMARGIN**<br>(training)<br>(n=631) | **ROCKET**<br>(training)<br>(n=304) | **KU Leuven**<br>(validation)<br>(n=3744) | **MH Hannover**<br>(validation)<br>(n=589) | **Necker Paris**<br>(validation)<br>(n=360) |
| --- | --- | --- | --- | --- | --- |
| Time after transplant (mo), median (IQR) | 12 (3-25) | 12 (3-44) | 12 (3-25) | 4 (2-12) | 12 (2-47) |
| Indicated biopsy, n (%) | 222 (35.2) | 134 (44.1) | 979 (26.1) | MD | MD |
| Pathologic diagnosis |  |  |  |  |  |
| ABMR, n (%) | 104 (16.5) | 107 (35.2) | 242 (6.7) | 36 (6.1) | 86 (23.9) |
| TCMR, n (%) | 82 (13.0) | 84 (27.6) | 665 (17.8) | 193 (33.3) | 47 (13.1) |
| Mixed ABMR/TCMR, n (%) | 28 (4.4) | 19 (6.2) | 79 (2.1) | 15 (2.5) | 13 (3.6) |
| BKVN, n (%) | 0 (0.0) | 13 (4.3) | 124 (3.3) | 23 (4.1) | 11 (3.1) |
| IFTA, n (%) | 210 (33.3) | 98 (32.9) | 780 (20.8) | 44 (8.2) | 188 (52.2) |
| Normal, n (%) | 312 (49.4) | 93 (30.6) | 2420 (65.9) | 317 (57.3) | 133 (36.9) |
| Laboratory test results at the time of the biopsy |  |  |  |  |  |
| Serum creatinine (μmol/L), median (IQR) | 150 (118-198) | 154 (119-208) | 141 (111-199) | 172 (131-234) | 176 (142-234) |
| DSA positivity, n (%) | 124 (19.7) | 87 (28.6) | 299 (8.3) | 11 (4.8) | 142 (41.0) |
| Proteinuria (g/L), median (IQR) | 0.10 (0.07-0.24) | 0.10 (0.07-0.34) | MD | 0.05 (0.04-0.10) | 0.20 (0.08-0.47) |

*Abbreviations: ABMR, active antibody-mediated rejection; BKVN, BK virus nephropathy; DSA, donor-specific antibodies; IFTA, interstitial fibrosis/tubular atrophy grade II; IQR, interquartile range; MD, missing data; Normal, refers to cases with no graft alterations; TCMR, T cell-mediated rejection.*

Detailed results of cross-validation in the training set are shown in the Supplemental Material. The ROC curves showed excellent performance with AUC of 0.99 (95% CI: 0.99-1.00), 0.98 (95% CI: 0.96-0.99) and 1.00 (CI 95%: 0.99-1.00) for ABMR, TCMR and IFTA classification, respectively. The calculated accuracy was 0.97, 0.95, 0.99 and 0.94 for ABMR, TCMR, IFTA and ABMR active/chronic active, respectively (arbitrary threshold set at 0.50). For the four models, the contribution (so-called “importance”) of the histological and clinical features is shown in Supplemental Figure 1.

Figure 2 shows the ROC and PR curves obtained in the three validation datasets. The ABMR algorithm yielded ROC curve AUC of 0.97 (95% CI: 0.92-1.00), 0.97 (95% CI: 0.96-0.97) and 0.95 (95% CI: 0.93-0.97), and PR curve AUC of 0.92, 0.72 and 0.84 for the Hannover, Leuven, and Necker datasets, respectively. In comparison, the minimum PR curve AUC for a No-Skill Classifier was 0.06, 0.07 and 0.24, respectively. For the TCMR model, the ROC AUCs were 0.94 (95% CI: 0.91-0.96), 0.94 (95% CI: 0.92-0.95) and 0.91 (95% CI: 0.86-0.95), the PR AUCs (minimum AUC for a No-Skill Classifier) were 0.91 (0.33), 0.83 (0.18) and 0.55 (0.13), respectively. For the IFTA model, the performance was even better with a minimum AUC of

**Figure 1:**
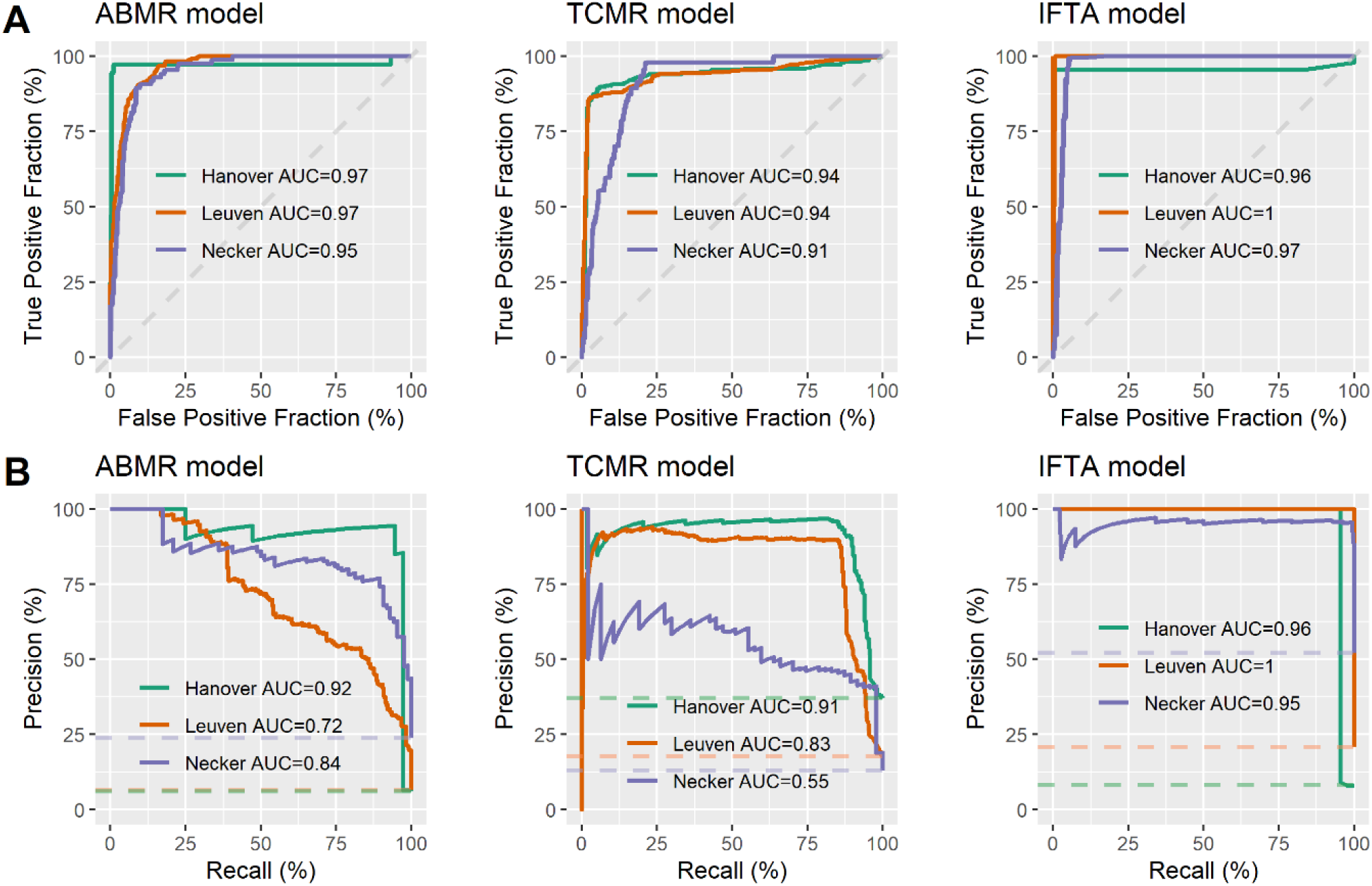
External validation of the Machine Learning estimators in three independent cohorts. (A) ROC curves of the ABMR model, the TCMR model and the IFTA model. (B) PR curves of the ABMR model, the TCMR model and the IFTA model. Abbreviations: ABMR, active antibody-mediated rejection; IFTA, interstitial fibrosis/tubular atrophy grade II; Precision, positive predictive value; Recall, sensitivity; TCMR, T cell-mediated rejection.

**Figure 2:**
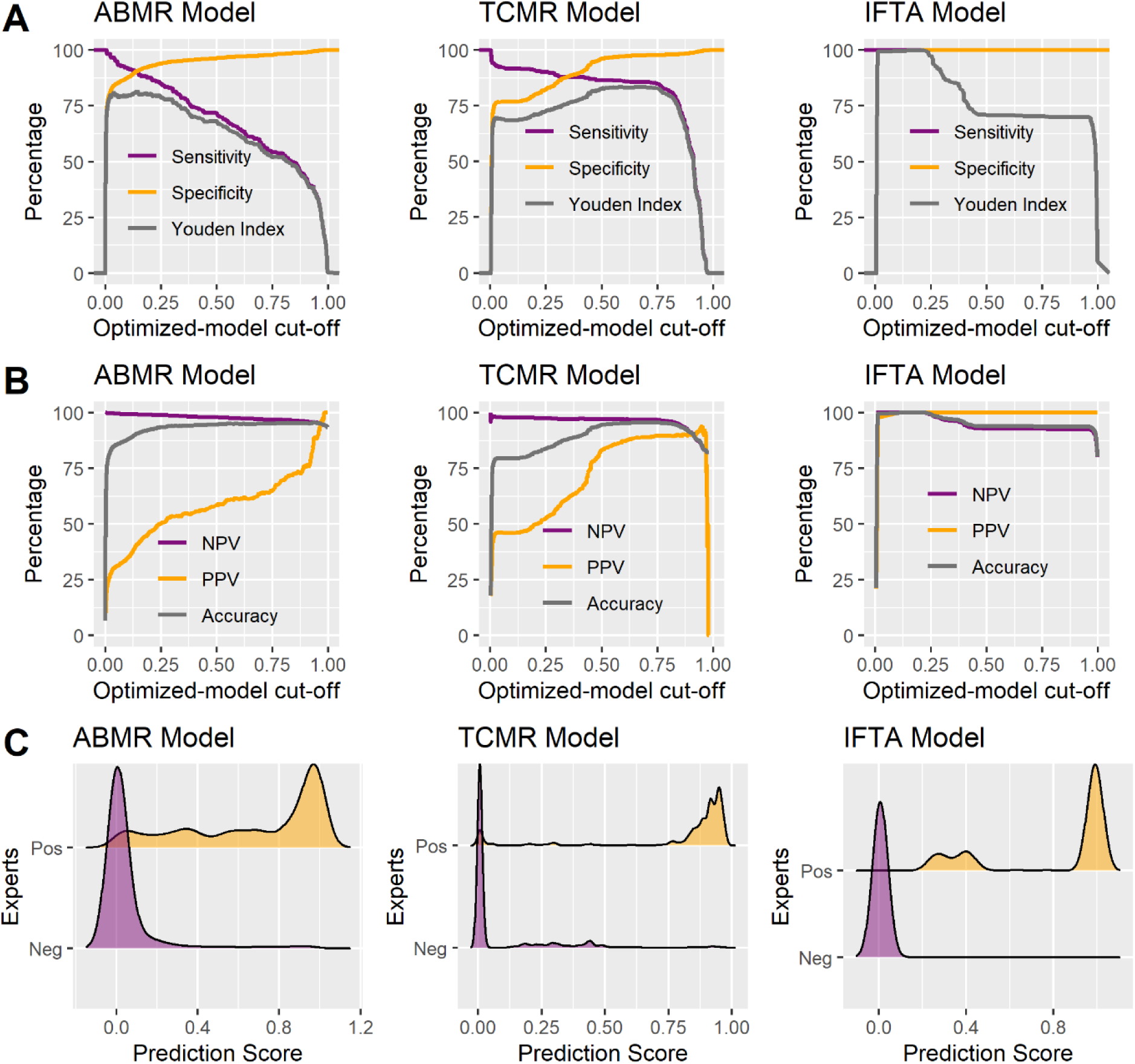
Choice of thresholds in the Leuven dataset. The plots at the bottom show the density of the scores. (A) Sensitivity, specificity, and Youden Index, depending on the cut-off for each model. (B) Negative predictive value, positive predictive value, and accuracy, depending on the cut-off for each model. (C) Density of the scores for each model. Abbreviations: ABMR, active antibody-mediated rejection; IFTA, interstitial fibrosis/tubular atrophy grade II; NPV, negative predictive value; PPV, positive predictive value; TCMR, T cell-mediated rejection; Youden Index, sensitivity + specificity - 1.

0.95 (95% CI: 0.90-1.00) for the ROC and PR curves, in all local datasets.

Thresholds were chosen to maximize accuracy in the Leuven cohort (Figure 2). We opted for a “grey zone” with two numerical cutoffs constituting its borders. The first cutoff was used to exclude each type of diagnosis with near certainty (to privilege sensitivity and negative predictive value), and the second to assert the diagnosis with similar near certainty (to privilege specificity and positive predictive value). The lower and upper thresholds were chosen at 0.10 and 0.75, respectively, for the binary models of ABMR and TCMR. Between these two thresholds, the ABMR grey zone includes 11.8, 0.6, and 2.1% of biopsies in the Leuven, Hannover, and Necker datasets, respectively. The TCMR grey zone includes 18.5, 1.1, and 0.9% of biopsies, respectively. For IFTA, the scores were already very well discriminated so we chose a unique threshold of 0.10. The features and performance of the final models are presented in Table 2.

**Table 2.** Thresholds chosen for, and performance of, the different algorithms.

|  |  | ABMR model |  | TCMR model |  | IFTA model |
| --- | --- | --- | --- | --- | --- | --- |
|  | Threshold | Low =<br>0.10 | High =<br>0.75 | Low =<br>0.10 | High =<br>0.75 | Unique =<br>0.10 |
| Leuven<br>dataset | Sensitivity | 91.7 | 54.1 | 91.7 | 84.8 | 100.0 |
|  | Specificity | 97.8 | 97.9 | 76.8 | 97.9 | 100.0 |
|  | NPV | 99.3 | 96.8 | 97.7 | 96.8 | 100.0 |
|  | PPV | 35.0 | 64.9 | 46.1 | 89.7 | 100.0 |
|  | % cases between the<br>two thresholds | 11.8 |  | 18.5 |  | NA |
| Hannover<br>dataset | Sensitivity | 97.2 | 91.7 | 90.2 | 82.3 | 95.5 |
|  | Specificity | 95.7 | 99.6 | 92.0 | 98.4 | 100.0 |
|  | NPV | 99.8 | 99.5 | 94.1 | 90.4 | 99.6 |
|  | PPV | 59.3 | 94.3 | 87.0 | 96.7 | 100.0 |
|  | % cases between the<br>two thresholds | 0.6 |  | 1.1 |  | NA |
| Necker<br>dataset | Sensitivity | 98.8 | 89.5 | 91.5 | 42.6 | 99.5 |
|  | Specificity | 65.0 | 90.9 | 80.5 | 91.8 | 94.8 |
|  | NPV | 99.4 | 96.5 | 98.4 | 91.8 | 99.4 |
|  | PPV | 47.0 | 75.5 | 41.3 | 62.5 | 95.4 |
|  | % cases between the<br>two thresholds | 2.1 |  | 1.9 |  | NA |

*Abbreviations: NA, not applicable; NPV, negative predictive value; PPV, positive predictive value.*

Table 3 presents the performance of the active/chronic active ABMR estimator in the Leuven cohort. The accuracy was 0.98 for biopsies above the upper cutoff of 0.75, as well as for biopsies above the lower cutoff of 0.10 (i.e., including the grey zone). The final accuracy of the combination of the two ABMR estimators (yes/no and active/chronic active) successively applied to the Leuven dataset was 0.95.

**Table 3.**
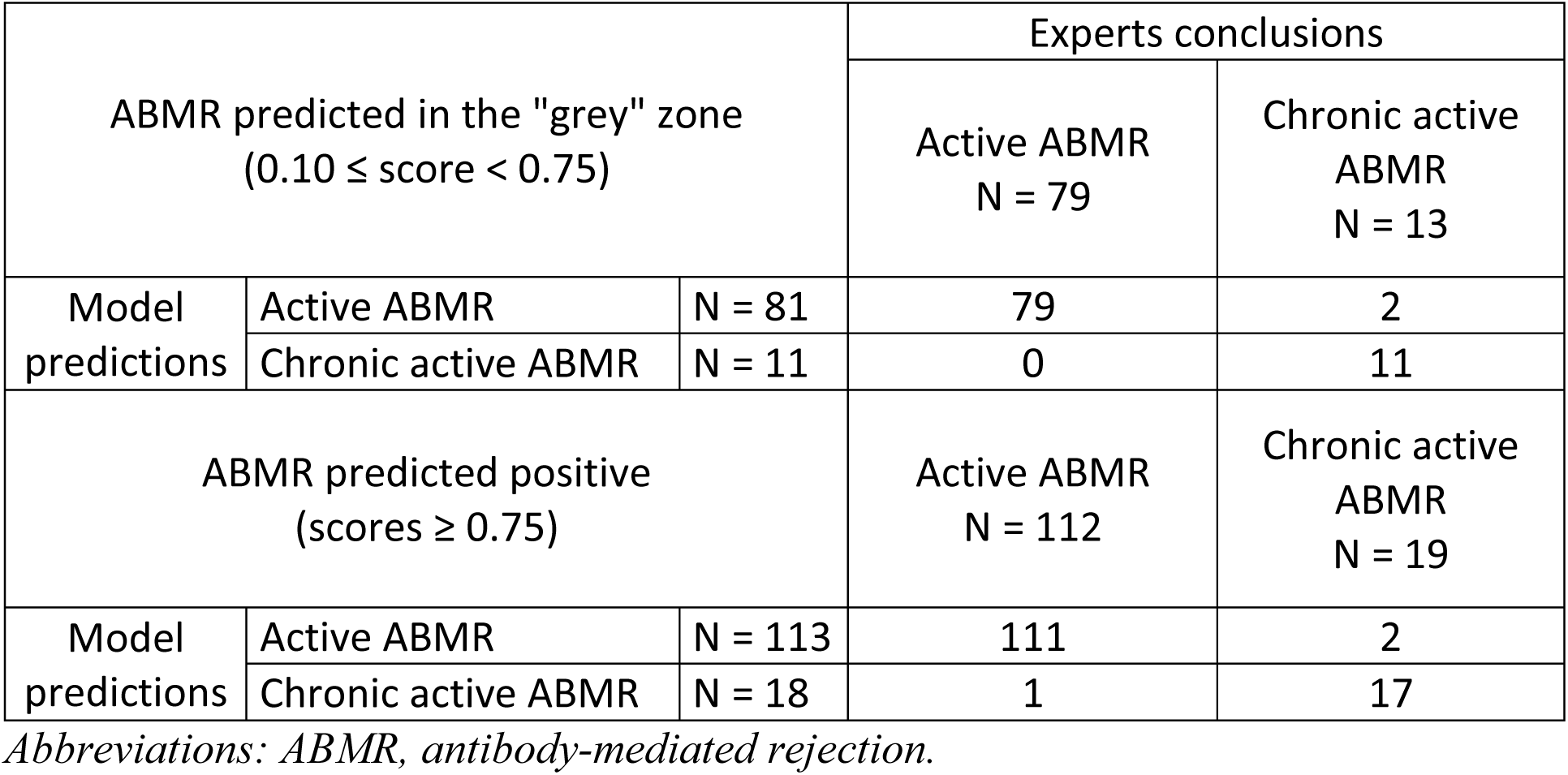
Evaluation of the ML estimations of active/chronic active ABMR as compared with expert conclusions in the Leuven dataset (n = 232)

Finally, we applied our algorithms to the 6 case-based scenarios used by Schinstock et al.^5^ for their international survey among clinicians and renal pathologists (to understand how the Banff ABMR classification is interpreted in practice). Model predictions were perfectly consistent with the reference diagnoses (100% agreement), without any doubt regarding the score values. Detailed input data and score results are presented in Supplemental Table 3.

Some specific cases were also studied in detail. Mixed ABMR/TCMR cases were predicted for 68%, 22% and 10% of them as ABMR, grey zone, and not ABMR, respectively. They were also independently classified for 85%, 7% and 8% of them as TCMR, grey zone and not TCMR respectively. Among the ABMR cases, 34%, 54% and 12% of those with negative DSA were classified as ABMR, the grey zone and not ABMR, respectively. Borderline TCMR^1^ cases were all predicted as TCMR.

## Discussion

Based on a training set made up of two large databases of kidney graft biopsy histological Banff scores and their combined pathological and clinical interpretation obtained in state-of-the art conditions, we developed AI algorithms able to automatically derive the main rejection diagnoses from the elementary Banff scores and a few clinical data. These algorithms showed excellent concordance with the clinical diagnoses made locally by specialized pathologists and transplant physicians in independent patient cohorts from three European transplant centers. Despite the fact that some scored biopsies had missing data in these different validation datasets (as is the case in usual practice), the performance of the estimators was still very good when up to 2 (and even sometimes 3 or 4) data per biopsy were missing. However, these missing values could not be accounted for in the experts’ annotation, which means that the reference diagnoses are uncertain and that the ML algorithms leading to the same conclusions do not provide proof of their actual performance.

Surprisingly, the predictors of each diagnosis retained by the algorithms after unsupervised selection were not all consistent with those proposed by the Banff classification. The IFTA grade can easily be assessed using only two criteria of the Banff classification, so it is not surprising that our model almost never failed. At least, this algorithm shows that no other criterion or clinical data influenced the expert decision for this phenotype. However, contrary to the Banff classification, the ct criterion was more important than ci (feature importance indicates how useful or valuable each feature is to a model). It is also worth noting that this was the only perfectly reproducible phenotype across hospitals and pathologists in our study. The reason why we could not predict the other phenotypes with ROC AUC = 1 despite the use of gradient boosting, an ensemble method literally based on decision trees, is probably due to the fact that the interpretation of the Banff classification for these phenotypes was not as reproducible among pathologists as that of IFTA.

In the TCMR model, i was much less used than t, whereas in the Banff classification they are of equal importance. Also, an increased v alone (≥ 1) triggers the diagnosis of TCMR in the Banff classification, whereas v was not part of the 8 most important variables in our model. Borderline TCMR was considered as TCMR in the learning and validation phases of the present study. Indeed, the aim was to propose a sensitive tool to detect rejection, considering the cost of false negative cases higher than that of false positives. Despite probably larger variability across centers for reporting borderline TCMR, consistency with the algorithm estimation was very high.

In the ABMR model (yes/no), cg was reported as the third most important variable, whereas the Banff classification uses this criterion only for the distinction between active and chronic active ABMR. Moreover, the v criterion was underused by the model (it is not one of the 8 most important variables) whereas it has the same importance as glomerulitis and peritubular capillaritis in the Banff classification. In addition, time after transplantation, serum creatinine and proteinuria had a higher rank than this v criterion among the 8 most important variables. The ABMR algorithm did not detect 12% of the ABMR cases without DSA. It is noteworthy that such cases are not consensually accepted as equivalent to those with positive DSA, since graft survival is not the same.^22^

The lower and upper thresholds were chosen at 0.10 and 0.75 for the binary models of ABMR and TCMR, respectively. Indeed, in the Leuven dataset used to set up these thresholds we observed that true negatives were uniformly distributed very close to the score of 0, whereas the scores of true positive cases were rather spread out between 0.75 and 1. Furthermore, we did not want to select a single best threshold value and overfit the Leuven dataset.

Grading elementary lesions is not always possible, because not all biopsies are deemed adequate. The number of glomeruli and arteries visible on the slides can be very small, making it impossible to assess all criteria. More generally, in the case of missing criteria, the algorithm performance might be reduced. For each classification algorithm, the measured importance of the variables involved (presented in supplemental data) points to the critical determinants. For instance, diagnosing ABMR requires at least the presence of the following data: g, ptc, cg. Therefore, for routine practice, an overlay of “if then else” rules should be applied upstream of the current algorithm to avoid making predictions in case one of these critical variables is missing. In contrast, the absence of one or a few minor predictors of ABMR, i.e. DSA, time after transplant, C4d staining, serum creatinine, and proteinuria, seems to be compatible with accurate prediction.

Interobserver reproducibility of kidney graft rejection diagnoses has been assessed many times in the past, sometimes limited to the detection and grading of the elementary lesions (the diagnoses being derived centrally using the Banff rules) while at other times encompassing the final diagnoses. For example, Marcussen et al.^2^ reported fair agreement for t, i and v, that were the only criteria used for grading the rejection at this period.^17^ The interobserver kappa score for grading the rejection severity was only 0.40 overall (fair agreement) while it was 0.56 when only the presence or absence of acute rejection was considered (moderate agreement). Furthermore, agreement was poor for the ah and g criteria, the latter being essential for the diagnosis of ABMR. The reproducibility of the elementary criteria (while the final phenotypes were assigned centrally) was also studied by Smith et al.^18^ The kappa score between pathologists on a diagnosis of ABMR (either active or chronic active) versus no rejection was better: mean (range) 0.70 (0.53-0.91). In this study, a “majority rules” approach was successfully used to reduce variability and increase kappa (from 0.70 to 0.82), similar to what had been done for the BIOMARGIN learning dataset used in the present study. In the study by Furness et al,^3^ the reference diagnosis of acute rejection was made based on the increase of serum creatinine in the week preceding the biopsy (or loss of the graft) with no other changes to explain such a change in creatinine. However, the criteria assessment was done blindly, meaning without taking into consideration any clinical feature. Once again, the interpretation of the elementary criteria was done automatically: only 74% and 47% of acute rejection episodes were detected, depending on whether the Banff “suspicious” grade was included or not. Gough^19^ and Veronese^20^ found moderate to good interobserver agreement in assigning a diagnosis of acute rejection. However, they did not mention whether the scores were interpreted in a centralized manner or by each pathologist individually. The inter-observer agreement about the conclusion drawn from the semi-quantitative criteria and the clinical context (as is done in routine practice) was not evaluated in any of these studies. Unfortunately, the vulnerability of the Banff classification to misinterpretations has already been demonstrated, especially for antibody-mediated rejection.^5^ Confirmation of the reliability of our algorithms was shown by their application to the 6 case-based scenarios used by Schinstock et al.^5^ for a large survey among clinicians and renal pathologists and their full agreement with the reference standards, as opposed to 26.1% and 35.5% differences for the pathologists and clinicians, respectively.

In retrospect, this study also points out the imperfect reproducibility of case classification within and across large European kidney transplantation centers. It also highlights how artificial intelligence can support the interpretation of the Banff elementary lesions, in order to help pathologists in their routine practice, as well as to minimize outcome uncertainty in multicenter clinical trials in kidney transplantation.

The main limitation of this approach is that it starts with human histological reading and elementary lesion grading of biopsies, i.e. on human skills and variability. However, the many AI tools being developed for digital image analysis may soon fill the gap^23–25^ and represent an alternative to time-consuming and non-reproducible visual scoring. For example, Hermsen et al. demonstrated the applicability of convolutional neural networks to automated histologic analysis of biopsy slides.^26^

Finally, artificial intelligence may help to standardize and facilitate the interpretation of complex clinical situations, such as those grouped under the terms “kidney graft rejection”. The algorithms described here can be adjusted to any future changes in the Banff criteria and diagnostic entities (such as chronic active TCMR as soon as an agreement has been reached). Biopsies of the learning data set will be re-examined by pathologist experts and new algorithms trained.

## Data Availability

The data that support the findings of this study are available on request from the corresponding author. The data are not publicly available due to privacy or ethical restrictions.

## Author contributions

M.L. and P.M. designed the study; J.-H. B., M.R. and P.K. carried out the examination of the biopsies; M.N., W.G and D.A assigned the reference diagnoses; M.L., P.M. and J.B.W. analyzed the data; M.L. and P.M. wrote the manuscript; M.N., W.G and D.A revised the manuscript. All authors approved the final version of the manuscript.

## Acknowledgments

This project was supported by ERACoSysMed-2, the ERA-Net for Systems Medicine in clinical research and medical practice (project ROCKET, JTC2 29).

## Supplemental material Table of Contents

Definitions of the phenotypes in the different external validation cohorts

- University Hospitals Leuven, Belgium
- Medizinische Hochschule Hannover, Germany
- University Hospital Necker Paris, France

Supplemental Table 1: Diagnostic characteristics.

Supplemental Table 2: XGBoost hyperparameters.

Supplemental Table 3: Machine-learning analysis of the 6 case-based scenarios used by Schinstock et al. for their survey among clinicians and renal pathologists.

Supplemental Figure 1: importance of the histological and clinical features for ML prediction.

Supplemental Figure 2: ROC curve analysis in the training dataset.

