## Supplemental material for "Machine learning-supported interpretation of kidney graft elementary lesions in combination with clinical data"

### Definitions of the phenotypes in the different external validation cohorts

#### University Hospitals Leuven, Belgium

In the Leuven cohort, a non-standard immunohistochemistry stain was used on frozen sections for C4d. The corresponding positivity threshold was 2/3. For the rest of the criteria, the Banff rules were strictly applied. For example, isolated v was considered as TCMR grade II, which is still under debate (Wohlfahrtova et al Clin Sci (Lond) 2018).

#### Medizinische Hochschule Hannover, Germany

In the Hannover cohort, the TCMR diagnosis was corrected when the sv40 staining was positive (to exclude false positives due to BK virus nephropathy -BKVN-). Indeed, in practice, when BK viremia is positive, pathologists systematically test for BKVN using SV40 staining (Hirsh et al Transplantation 2005). In this cohort, all the BKVN cases had positive BK viremia.

#### University Hospital Necker Paris, France

In the Necker Cohort, the Banff rules were strictly applied in most cases, except for adjustments made by the clinicians in complex situations. In this center too, SV40 staining was performed in case of concomitant BK viremia.

### Supplemental Table 1: Diagnostic characteristics.

| Variables | Normal (BIOMARGIN) N = 312 | ABMR (BIOMARGIN) N = 104 | TCMR (BIOMARGIN) N = 82 | IFTA (BIOMARGIN) N = 210 | Missing data (BIOMARGIN) N = 631 (100%) | Active ABMR (ROCKET) N = 63 | Chronic active ABMR (ROCKET) N = 44 |
| --- | --- | --- | --- | --- | --- | --- | --- |
| g, median (IQR) | 0 (0-0) | 1 (1-2) | 0 (0-1) | 0 (0-0) | 0 (0.0%) | 1 (1-2) | 2 (1-2) |
| ptc, median (IQR) | 0 (0-0) | 2 (1-2) | 0 (0-1) | 0 (0-0) | 1 (0.2%) | 2 (1-2) | 1 (1-2) |
| v, median (IQR) | 0 (0-0) | 0 (0-0) | 0 (0-0) | 0 (0-0) | 21 (3.3%) | 0 (0-0) | 0 (0-0) |
| C4d, median (IQR) | 0 (0-0) | 0 (0-2) | 0 (0-0) | 0 (0-0) | 21 (3.3%) | 0 (0-2) | 0 (0-1) |
| DSA positivity (%) | 39 (13.2) | 56 (54.9) | 30 (37) | 33 (16.4) | 27 (4.3%) | 35 (55.6) | 21 (47.7) |
| cg, median (IQR) | 0 (0-0) | 0 (0-2) | 0 (0-0) | 0 (0-0) | 6 (1.0%) | 0 (0-0) | 3 (2-3) |
| i, median (IQR) | 0 (0-0) | 0 (0-0) | 1 (0-2) | 0 (0-0) | 1 (0.2%) | 0 (0-1) | 0 (0-0) |
| t, median (IQR) | 0 (0-0) | 0 (0-1) | 1 (1-2) | 0 (0-0) | 1 (0.2%) | 0 (0-1) | 0 (0-1) |
| ti, median (IQR) | 0 (0-0) | 0 (0-1) | 1 (1-3) | 0 (0-1) | 9 (1.4%) | 0 (0-1) | 0 (0-1) |
| ct, median (IQR) | 0 (0-1) | 1 (0-2) | 1 (0-2) | 2 (2-3) | 1 (0.2%) | 0 (0-1) | 2 (1-3) |
| ci, median (IQR) | 0 (0-1) | 1 (0-2) | 1 (0-2) | 2 (2-3) | 1 (0.2%) | 0 (0-2) | 2 (1-3) |
| ah, median (IQR) | 0 (0-1) | 1 (0-3) | 0 (0-1) | 1 (0-2) | 9 (1.4%) | 0 (0-1) | 2 (1-3) |
| cv, median (IQR) | 0 (0-1) | 1 (0-3) | 1 (0-2) | 2 (1-2) | 33 (5.2%) | 0 (0-2) | 2 (1-3) |
| Serum creatinine (µmol/L), median (IQR) | 138 (110-169) | 188 (149-240) | 185 (131-254) | 169 (133-229) | 0 (0.0%) | 183 (127-237) | 198 (156-254) |
| Proteinuria (g/L), median (IQR) | 0.09 (0.07-0.15) | 0.30 (0.07-1.13) | 0.11 (0.07-0.32) | 0.11 (0.07-0.30) | 0 (0.0%) | 0.13 (0.07-0.51) | 1.19 (0.25-2.29) |
| Time after transplant (mo), median (IQR) | 3 (3-12) | 32(5-120) | 12 (3-53) | 24 (12-60) | 0 (0.0%) | 11 (3-25) | 125 (47-186) |

*Abbreviations: ABMR, active antibody‐mediated rejection; ah, arteriolar hyalinosis criterion; c4d, linear C4d staining in ptc or medullary vasa recta criterion; cg, chronic transplant glomerulopathy criterion; ci, interstitial fibrosis in cortex criterion; ct, tubular atrophy in cortex criterion; cv, arterial intimal fibrosis criterion (fibrointimal thickening); DSA, donor-specific antibodies; g, glomerulitis criterion; i, inflammation in non‐scarred cortex criterion; IFTA, interstitial fibrosis/tubular atrophy grade II; IQR, interquartile range; Normal, refers to cases with no graft alterations; ptc, peritubular capillaritis criterion; t, tubulitis in cortical tubules within non‐scarred cortex criterion; TCMR, T cell-mediated rejection; ti, total cortical inflammation criterion; v, endarteritis (intimal arteritis).*

### Supplemental Table 2: XGBoost hyperparameters.

|  | Model | | | |
| --- | --- | --- | --- | --- |
| Hyperparameters | Active ABMR | TCMR | IFTA | ABMR active/chronic active |
| mtry | 2 | 20 | 15 | 13 |
| Trees | 1000 | 1000 | 1000 | 1000 |
| min_n | 2 | 6 | 4 | 2 |
| Tree depth | 3 | 9 | 4 | 2 |
| Learning rate | 0.024 | 0.015 | 0.014 | 0.017 |

*Abbreviations: Learning rate, rate at which the boosting algorithm adapts from iteration-to-iteration; min_n, minimum number of data points in a node that are required for the node to be split further; mtry, number of predictors that will be randomly sampled at each split when creating the tree models; Tree depth, maximum depth of the tree (i.e., number of splits); Trees, number of trees contained in the boosted ensemble (i.e., number of boosting iterations).*

### Supplemental Table 3: Machine-learning analysis of the 6 case-based scenarios used by Schinstock et al. for their international survey among clinicians and renal pathologists.

| **Case-based scenarios** | | | | | | | | | | | | | | | | | | **Machine-learning interpretation** | | | | | | | |
| --- | --- | --- | --- | --- | --- | --- | --- | --- | --- | --- | --- | --- | --- | --- | --- | --- | --- | --- | --- | --- | --- | --- | --- | --- | --- |
| Case # | g | ptc | c4d | cg | v | DSA | i | t | ti | ct | ci | ah | cv | Serum creatinine (µmol/L) | Proteinuria | Time between transplantation and biopsy (months) | Reference diagnosis | ABMR score | Active ABMR prediction | Chronic active ABMR score | Chronic active ABMR prediction | TCMR score | TCMR prediction | IFTA score | IFTA prediction |
| 1 | 2 | 1 | 1 | 1 | MD | 1 | 0 | 0 | MD | 0 | 0 | MD | MD | 194.52 | MD | 48 | Chronic active ABMR | 0.99 | positive | 0.84 | positive | 0.01 | negative | 0.00 | negative |
| 2 | 1 | 2 | 0 | MD | MD | 1 | 0 | 0 | MD | 0 | MD | MD | MD | 123.79 | MD | 36 | Acute/active ABMR | 0.98 | positive | 0.04 | negative | 0.02 | negative | 0.01 | negative |
| 3 | 1 | 2 | 0 | 1 | MD | 1 | 0 | 0 | MD | 0 | 0 | MD | MD | 194.52 | MD | 120 | Chronic active ABMR | 1.00 | positive | 0.85 | positive | 0.01 | negative | 0.00 | negative |
| 4 | 2 | 2 | 0 | MD | MD | 0 | 0 | 0 | MD | 0 | 0 | MD | MD | 150.31 | MD | 12 | Histologic features of ABMR without detectable anti-HLA antibody | 0.96 | positive | 0.05 | negative | 0.02 | negative | 0.01 | negative |
| 5 | 2 | 2 | 0 | MD | MD | 1 | 0 | 0 | MD | 0 | 0 | MD | MD | 150.31 | MD | 6 | Acute/active ABMR | 0.99 | positive | 0.05 | negative | 0.01 | negative | 0.01 | negative |
| 6 | 1 | 2 | 0 | 0 | MD | 1 | 2 | 3 | MD | MD | MD | MD | MD | 221.05 | MD | 18 | Mixed acute T cell mediated rejection and ABMR | 0.98 | positive | 0.01 | negative | 0.98 | positive | 0.00 | negative |

*Abbreviations: ah, arteriolar hyalinosis criterion; c4d, linear C4d staining in ptc or medullary vasa recta criterion; cg, chronic transplant glomerulopathy criterion; ci, interstitial fibrosis in cortex criterion; ct, tubular atrophy in cortex criterion; cv, arterial intimal fibrosis criterion (fibrointimal thickening); dsa, donor-specific antibodies; g, glomerulitis criterion; i, inflammation in non‐scarred cortex criterion; MD, missing data; ptc, peritubular capillaritis criterion; t, tubulitis in cortical tubules within non‐scarred cortex criterion; ti, total cortical inflammation criterion.*

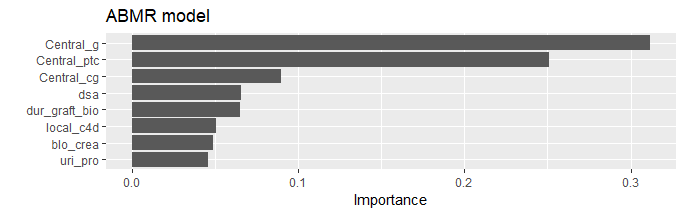

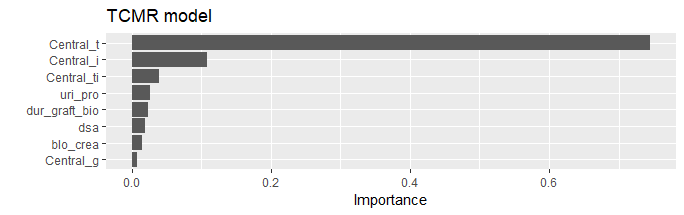

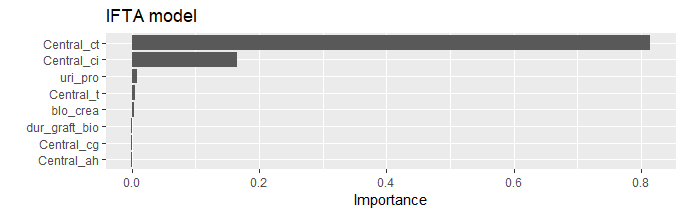

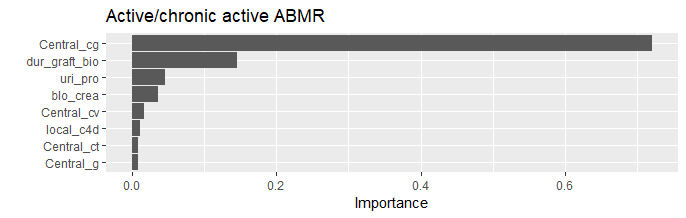

### Supplemental Figure 1: importance of the histological and clinical features for ML predictions of ABMR, TCMR, IFTA and active/chronic active ABMR.

“Importance” provides a score that indicates how useful or valuable each feature was in the construction of the boosted decision trees within the model. The more an attribute is used to make key decisions with decision trees, the higher its relative importance.

*Abbreviations: blo_crea, serum creatinine; Central_ah, arteriolar hyalinosis criterion; Central cg, chronic transplant glomerulopathy criterion; Central_ci, interstitial fibrosis in cortex criterion; Central_ct, tubular atrophy in cortex criterion; Central_cv, arterial intimal fibrosis criterion (fibrointimal thickening); Central_g, glomerulitis criterion; Central_i, inflammation in non‐scarred cortex criterion; Central ptc, peritubular capillaritis criterion; Central_t, tubulitis in cortical tubules within non‐scarred cortex criterion; Central_ti, total cortical inflammation criterion; dsa, donor-specific antibodies; dur_graft_bio, time after transplant; local_c4d, linear C4d staining in ptc or medullary vasa recta criterion; uri_pro, proteinuria.*

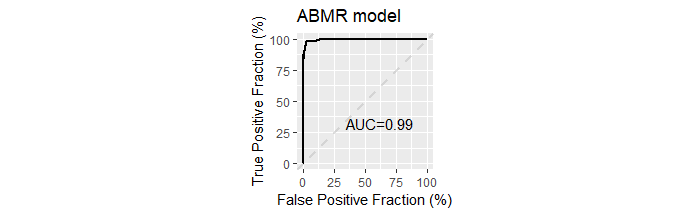

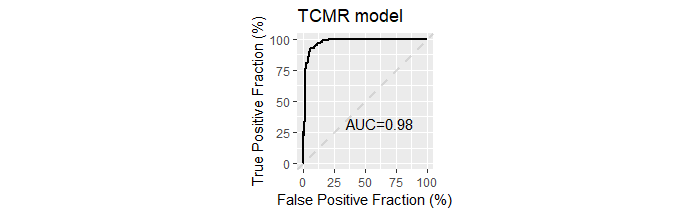

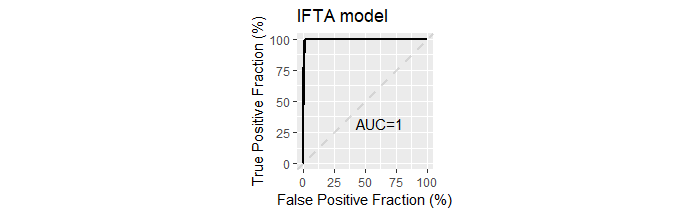

### Supplemental Figure 2: ROC curve analysis in the training dataset.

The accuracy was 0.97, 0.95, 0.99 and 0.94 for the ABMR model, the TCMR model, the IFTA model and the ABMR active/chronic model, respectively (with thresholds arbitrarily set at 0.50).
